# Pioglitazone for optimisation of clinical nutrition therapy in the critically ill patient: A Systematic Review

**DOI:** 10.1101/2022.06.07.22276074

**Authors:** Thomas J McClelland, Alexander J Fowler, Thomas Davies, Rupert Pearse, John Prowle, Zudin Puthucheary

## Abstract

**Background:** Skeletal muscle wasting is a major determinant of physical functional disability in critical illness survivors, and contributes to post-intensive care syndrome. As yet, no therapies exist to address this major public health issue. Intramuscular bioenergetic failure and inflammation are understood to be the underpinning mechanisms, and carbohydrate and lipid oxidation are impaired. This systematic review synthesises the evidence that peroxisome proliferator-activated receptor gamma agonists may be a therapeutic option to optimise clinical nutrition in critically ill patients.

**Method:** Systematic review and meta-analysis.

**Results:** Fourteen studies over 19 publications were included. Lean body mass was unaffected (n=174). Pioglitazone treatment resulted in periperal insulin sensitivity increasing 30-71% (Standardised mean change 0.97 (95%CI 0.36-1.58; n=213). Intramuscular Tumour Necrosis Factor Alpha concentrations decreased in treatement arms (n=29) as did circulating interlukin-6 and Tumour Necrosis Factor Alpha (n=53). Intramyocellular Lipid concentrations decreased by 34-40% with pioglitazone therapy (n=60). Treatment increased intramuscular markers of Oxidative Phosphorylation (n=55), mitochondrial biogenesis(PGC1α and PGC1β; n=26) and β-oxidation (n=29)

**Conclusions:** Pioglitazone therapy increases skeletal muscle insulin sensitivity, decreases intramyocellular lipid accumulation and systemic and intramuscular inflammation. Where lean body mass was measured, this was seen to increase. Pioglitazone may be an adjunctive therapy to optimise clinical nutrition in acutely unwell patients.

**Clinical relevancy statement:** Pioglitazone may optimise clinical nutrition therapy in critically ill patients by normalising carbohydrate and lipid metabolism.

## Background

Skeletal muscle wasting is a major determinant of physical functional disability in critical illness survivors and contributes to post-intensive care syndrome^1,2^. As yet, no therapies exist to address this major public health issue^3^. Muscle wasting in critical illness is the result of decreased muscle protein synthesis, which is underpinned by cellular bioenergetic failure and intramuscular inflammation^1,4^. These specific pathophysiological processes impair the efficacy of the usual stimulants of muscle protein synthesis: resistance exercise and amino acids^5^.

Cellular bioenergetics are adversely affected by impaired glucose metabolism and oxidation, upregulation of lipolysis, and mitochondrial dysfunction due to imbalance of reactive oxygen species^6^. The abnormalities of metabolism observed on a cellular and systemic level in critical illness share common features with metabolic syndrome. Key areas of pathophysiological overlap include: hyperglycaemia secondary to global insulin resistance, dysregulation of adiponectin, dyslipidaemia and pro-inflammatory response mediated in part by interleukins and Tumour Necrosis Factor Alpha (TNF-α)^7^.

Therapeutic targeting of Peroxisome proliferator-activated receptor gamma (PPARγ), a nuclear hormone receptor involved in the regulation of glucose and lipid metabolism, has been used in the treatment of 2 diabetes mellitus and metabolic syndrome through synthetic insulin sensitisers such as thiazolidinediones^8,9^. PPARγ has a central role in adipogenesis, regulation of fatty acid metabolism, and mediation of peripheral insulin sensitivity via adipocytokine secretion^10^. In skeletal muscle cells, PPARγ agonism has been shown to provide protection against lipotoxicity through mobilisation of fatty acids, enhance insulin signalling and insulin-mediated glucose uptake, and improve cell bioenergetics through promotion of normal mitochondrial function^11,12^. The combination of these metabolic modulatory effects would potentially restore substrate utilisation capacity enabling nutritional carbohydrates and lipids to ameliorate bioenergetics failure. This in turn would optimise the anabolic effects of nutritional amino acids and exercise, minimising muscle mass loss and improving patient outcomes.

The use of PPARγ agonism in critical illness has not been previously investigated. While there are numerous potential therapeutic benefits of PPARγ agonism in critical illness, the adverse effects of thiazolidinediones include peripheral oedema and congestive cardiac failure may limit tolerance in critically ill patients, though systematic reviews of randomised controlled trials suggest an excellent safety profile^8,13^. The objectives of this review are to examine the metabolic effects of PPARγ agonism in randomised controlled trials and specifically synthesise the evidence on this impact on skeletal muscles metabolism and function.

## Methods

The study protocol was registered on Friday 22^nd^ January 2022 prior to starting our review with the PROSPERO international prospective register of systematic reviews (CRD42022289052) and is reported in line with the Preferred Reporting Items for Systematic Reviews and Meta-Analyses (PRISMA) guidelines^14^.

### Information Sources and Eligibility Criteria

We electronically searched MEDLINE (via Ovid), CENTRAL, EMBASE (via Ovid), and CINAHL (via HDAS) as well as trial registries; ClinicalTrials.gov and the WHO International Clinical Trial Registry Platform (ICTRP) from inception to 23^rd^ January 2022 for original articles in peer reviewed journals excluding conference proceedings, preprints and publications in abstract form only. Terms were combined with Boolean operators to identify randomised controlled trials and quasi-randomised controlled trials in adults (age >18years), with no language restrictions. We included trials in which pioglitazone was administered, with no restriction on dose or dosing regimen. We excluded case studies as well as reviews, commentaries, opinion-pieces, preprints, unpublished studies, editorials and conference abstracts without subsequent study publication.

### Outcomes and measures of effect

The primary outcomes were physical function and symptoms, muscle mass and function, or body composition and muscular compositional change. The measures of effects for physical function and symptoms were: functional status using validated performance scales or exercise testing (including timed up and go testing, six-minute walk test, cardiopulmonary exercise testing). For muscle mass and function, the measures of effect were cross sectional area of femoral/quadriceps, muscle layer thickness, hand grip strength, leg press power, aerobic exercise or cardiovascular fitness. For body/muscle composition change, effect was assessed by a measure of change in lean tissue mass (DXA) or levels of intramyocellular lipid (IMCL).

Secondary outcomes included: muscle insulin sensitivity (glucose uptake measurements); muscle gene expression of interest (e.g. TNF-α, TIMP-3, TACE, oxidative enzymes, GLUT-4); muscle mitochondrial effects (measures of metabolic flexibility, maximal ATP synthetic capacity (ATPmax), mitochondrial copy number, mitochondrial proteome) and activity or expression of markers of intra-muscular inflammation e.g. super-oxidase dismutase 2, p-ERK, JNK.

### Search strategy

The full search strategies performed are listed in supplementary tables 1-3. After the first round of record screening and full text eligibility for each search, additional potentially relevant terms were extracted from short-listed papers, and further searches were performed to capture any other relevant records. In addition, snowball methods, pursuing references of references and electronic citation tracking, were used as is recommended for reviews of complex evidence.^15^

### Selection process

Studies resulting from the search strategy from each bibliographic database were screened for duplication. Two review authors (TJM, ZP) independently screened all records (title and abstract) for inclusion against pre-specified inclusion criteria (adults aged 18 or over, pioglitazone therapy administered, muscle-related outcome measure reported, randomised controlled trials). Any disagreements were resolved through independent review by a third author (JP).

Full text study reports/publications were retrieved for studies resulting from the initial abstract screening. Two authors (TJM, ZP) screened full text articles against the inclusion and exclusion criteria. Reason for exclusion were documented for ineligible studies. Any disagreements were resolved through independent review by a third author (JP).

### Data extraction

Data were extracted from each study using a data extraction form by one author (TJM), which included:

- Methodology (study design, context, patient selection, randomisation protocol, study length, date of study)
- Characteristics of study participants (patient demographics, baseline characteristics, disease studied, inclusion and exclusion criteria)
- Details of intervention used (dose, dosing regime, length of administration)
- Details of comparator arm (placebo/sham therapy, concomitant medications)
- Outcomes (outcomes investigated, measures of effect, secondary outcomes)
- Additional details for risk of bias assessment (pre-specified analysis plan, missing data, study funding, conflicts of interest)

Where direct data extraction was not possible, either study authors were contacted for the requisite data or online data extraction software was used to extract data from study plots^16^. Extracted data were spot-checked independently by another author (ZP). Any disagreement was resolved by consensus or by a third author (JP).

### Risk of bias assessment

Two authors (TJM, ZP) performed risk of bias assessment for each study reporting the primary outcomes of interest using the Cochrane risk-of-bias tool for randomised trials (RoB 2)^17^.

Studies were categorised systematically using the RoB2 tool into ‘Low’ risk of bias, ‘High’ risk of bias or ‘some concerns’. Any disagreement in categorisation were resolved through consensus or through an independent reviewer (JP).

### Data synthesis

Studies with common methodology for each outcome measure were grouped to facilitate comparability. Continuous data outcomes measures previously specified were collected for quantitative analysis. Data are expressed as mean (SD), median (IQR), or number (%) where appropriate. Where sufficient data exists to conduct meta-analysis, this was performed using the ‘meta’ package for R statistical software ^18,19^. Given the broad inclusion criteria, a random-effects model with an appropriate summary measure was used. Where heterogeneity was evaluated using t^2^ and I^2^ statistics.

There was sufficient data to conduct meta-analysis for peripheral insulin sensitivity – measured by ‘glucose disposal rate’ (R_d_). R_d_ is a continuous variable and, where available, mean baseline and endpoint outcome values for each intervention arm were extracted from studies reporting R_d_. Variance measures were standardised to standard deviation. Given the included studies did not report R_d_ in directly comparable units, baseline and endpoint means were used to calculate a mean change between the time points. No study reported mean change or variance of the mean change in either control or intervention arms. To calculate the standard deviation of change, the following equation was used^20^:

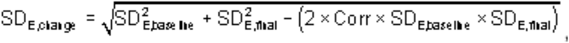

“Corr” represents the correlation co-efficient. This was calculated from raw data kindly submitted to the review team by Glintborg using the following equation^20,21^:

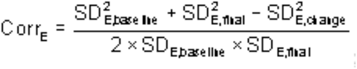

A correlation co-efficient was calculated for both control (0.881) and intervention (0.998) arms. A sensitivity analysis using these two correlation coefficients was conducted. One study did not report baseline values^22^. This was not included in the meta-analysis of mean change. A meta-analysis using an inverse variance model based on standardised mean change for the pooled results is reported.

## Results

A total of 2032 publications were identified by the search strategy. Following removal of duplicates (n=576), 1456 records were screened from which 57 full-text documents were assessed against the inclusion and exclusion criteria. In total, 14 studies (19 publications) met inclusion criteria as per Figure 1, and are detailed in Tables 1 and 2. All studies were single-centre. Study dose of pioglitazone varied from 15-45mg once a day for 12-26 weeks. The most common dose was 45 mg (10/15 studies).

**Figure 1.**
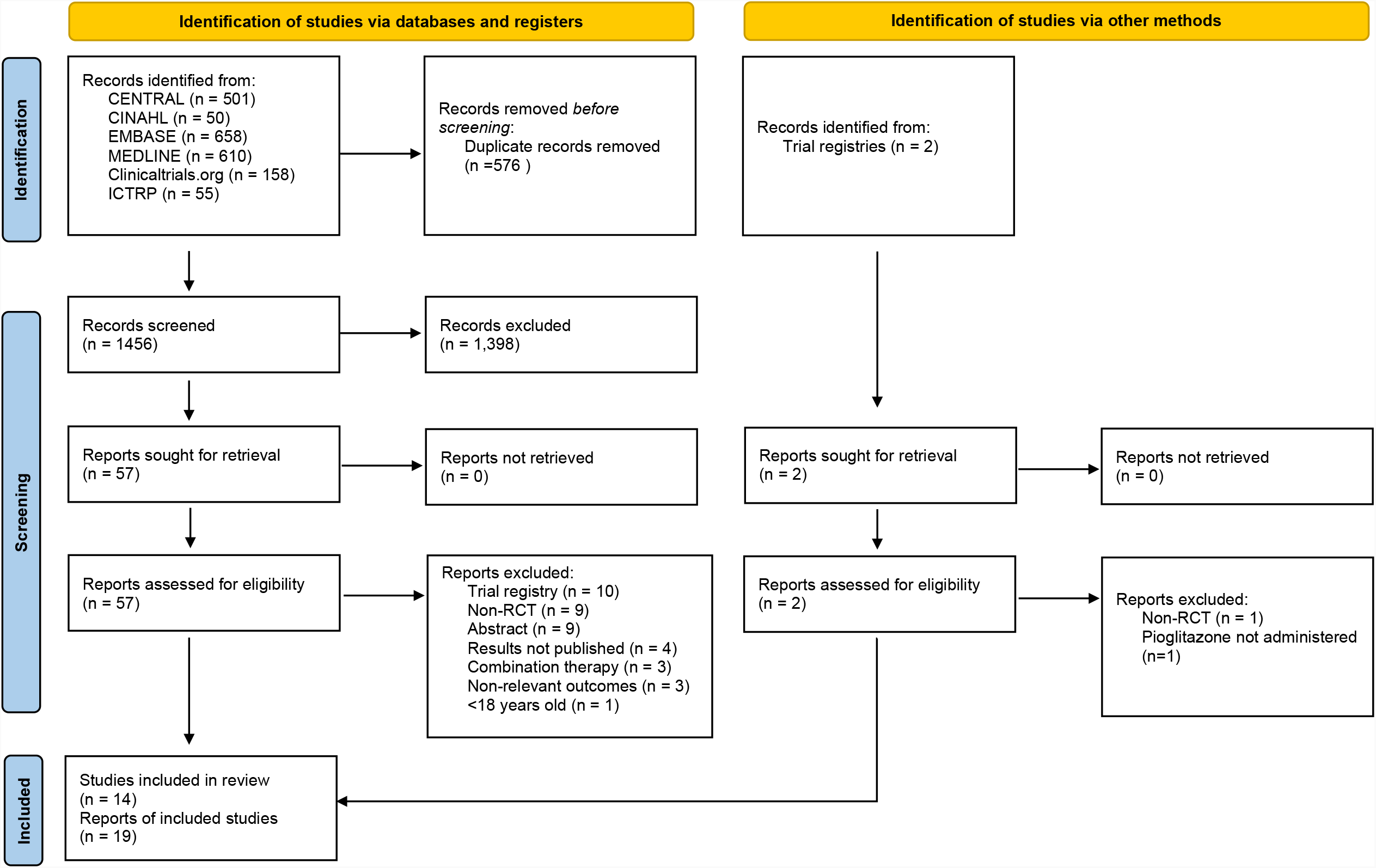

**Table 1:** Studies extracted with data pertaining to the primary outcomes of physical function and symptoms, muscle mass and function, or body composition and muscular compositional change. BMI= Body Mass Index, DEXA= Dual-energy X-ray absorptiometry.

| Study | Quality | Population | Intervention | Comparison | Duration | Outcomes |
| --- | --- | --- | --- | --- | --- | --- |
| Finlin<sup>25</sup> | Randomised<br>Open-label | BMI >27, pre-diabetes or metabolic syndrome (n=26) | 30mg/day Pioglitazone (n=13, n=12 analysed) | Combination therapy of 30mg/day pioglitazone and mirabegron 50mg/day (n=13) | 12 weeks | No change in total lean mass (DEXA, Kg) between intervention arms (Pioglitazone, p=0.73, combination therapy, p=0.76) |
| Shea<sup>23</sup> | Randomised | Older (65-79 years) subjects, non-diabetic (n=88) | 15mg/day [first 3 weeks], then 30mg/day Pioglitazone (n=44, n=41 analysed) | No pioglitazone (n=44, n=40 analysed) | 16 weeks | No change in total lean mass (DEXA, Kg) from baseline (male or female)<br>No change in muscle attenuation (HU)<br>No change from baseline (male or female) |
| Marsh<sup>24</sup> |  |  |  |  |  | No difference in mean leg press power (Watts),<br>Increase in women randomised to pioglitazone plus resistance training (p=0.006)<br>No change in baseline body mass (Women, p=0.22. Men, p=0.25 between group difference) |
| Shah<sup>27</sup> | Randomised<br>Placebo-controlled | Insulin-dependent Type II diabetic, obese (n=25) | Insulin + Pioglitazone (45mg/day) (n=12) | Insulin + placebo (n=13) | 12-16 weeks | Increase total lean body mass (DEXA, kg) in both intervention arms (+1.29 ± 0.61 kg with intensive insulin treatment alone (p = 0.057) and +1.92 ± 0.74 kg with added pioglitazone (p = 0.02) |
| Zanchi<sup>22</sup> | Double-blind<br>Randomised<br>Cross-over | Non-diabetic, end-stage renal disease (haemodialysis (n=8), peritoneal dialysis (n=4)) | 45mg/day (n=10 analysed) | Placebo (n=9 analysed) | 16 weeks | No change in lean mass (DEXA, Kg) |
| Rasouli<sup>51</sup> | Randomised | Generally healthy subjects with impaired glucose tolerance (n=23) | Two week dose escalation to max dose of 45mg/day (n=11) | Metformin (1000mg twice daily) | 10 weeks | Increase in lean body mass (DEXA, Kg) in pioglitazone group (p<0.005) |

**Table 2:**
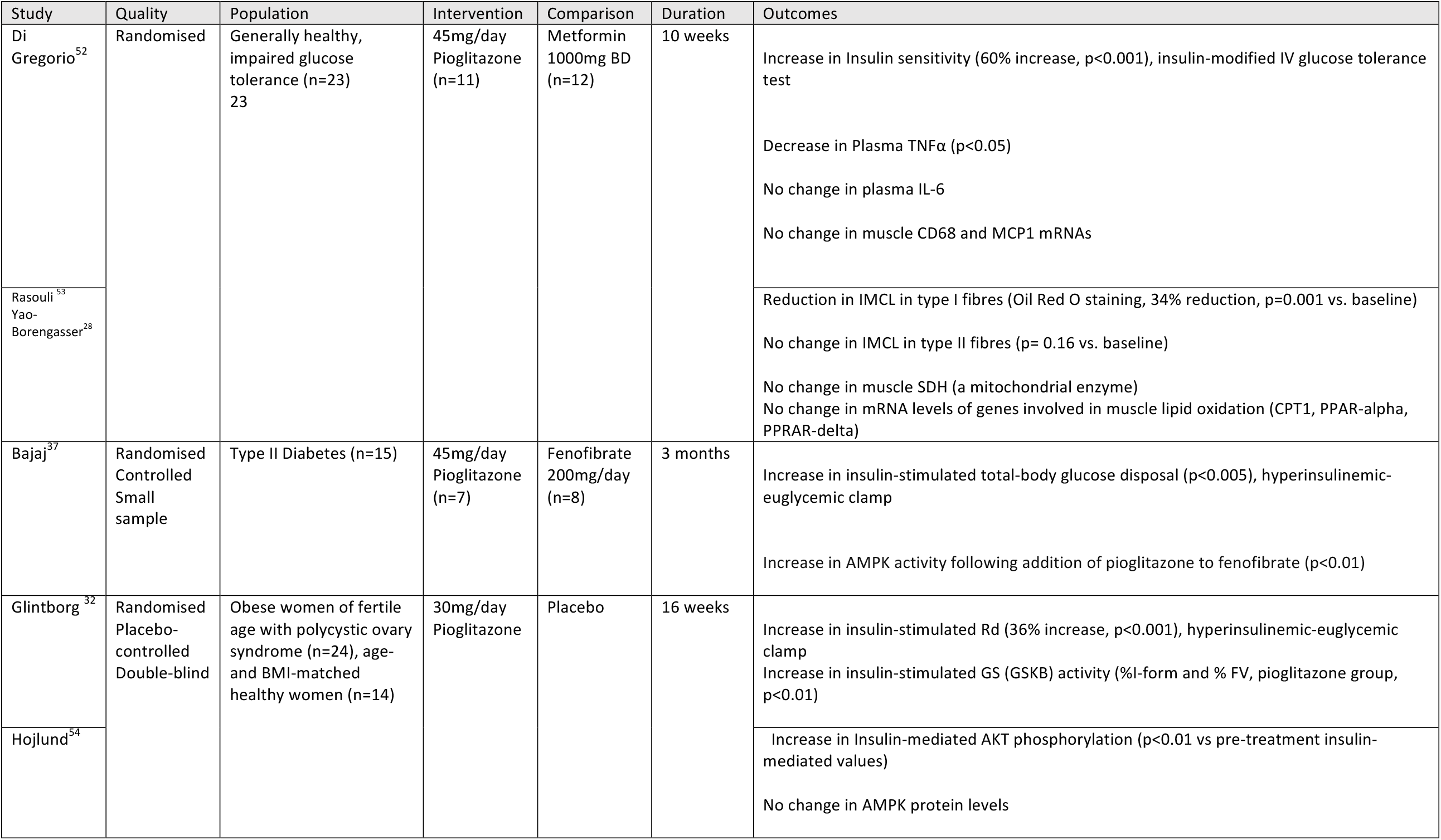
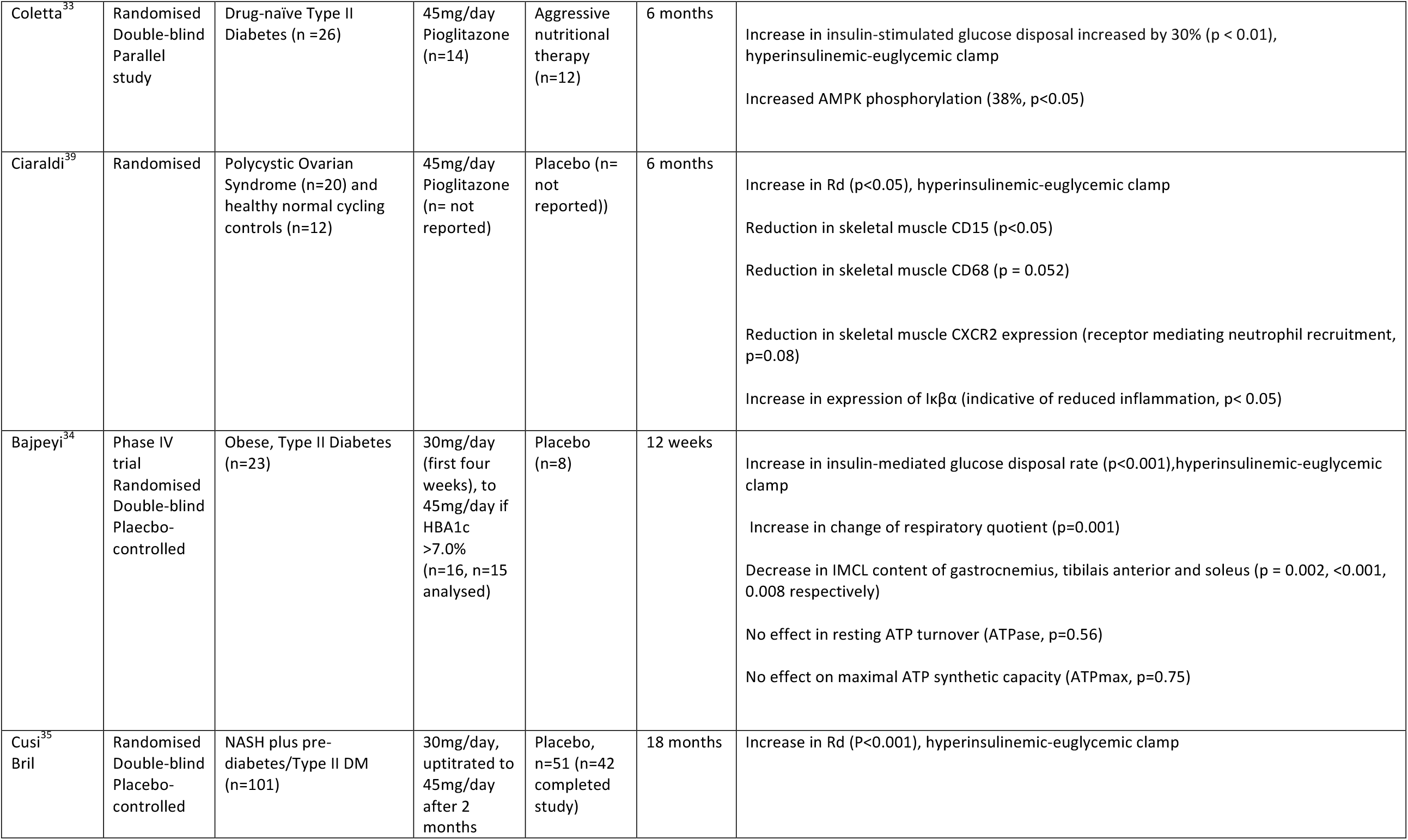
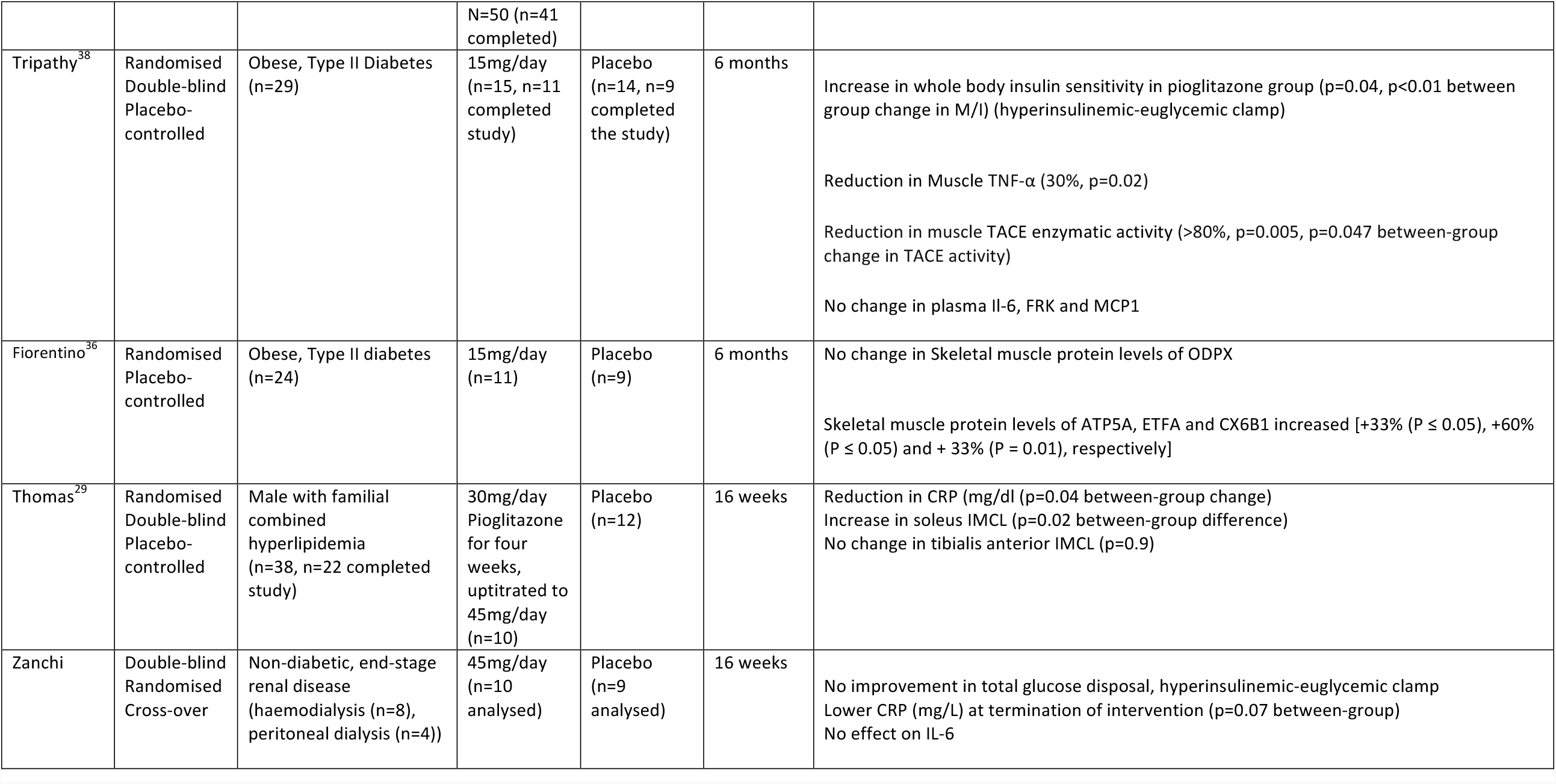
Studies extracted with data pertaining to secondary outcomes. BMI= Body Mass Index TNFα= Tumour Necrosis Factor alpha; MCP1= Macrophage Chemoattractant Protein 1; pGSK3β= phosphorylated Glycogen Synthesase Kinase 3-beta; PI3K= Phosphoinositide 3-kinase; pAKt= phosphorylated protein kinase B; Iκβα= Nuclear Factor Kappa Beta target gene; IMCL= Intramyocellular lipids; AMPK= Adenosine MonoPhosphate Kinase, PGC1 α β= Peroxisome proliferator-activated receptor gamma co-activator alpha and beta; ATP= Adenosine TriPhosphate.

| Study | Quality | Population | Intervention | Comparison | Duration | Outcomes |
| --- | --- | --- | --- | --- | --- | --- |
| Di Gregorio<sup>52</sup> | Randomised | Generally healthy, impaired glucose tolerance (n=23)<br>23 | 45mg/day<br>Pioglitazone<br>(n=11) | Metformin<br>1000mg BD<br>(n=12) | 10 weeks | Increase in Insulin sensitivity (60% increase, $p<0.001$), insulin-modified IV glucose tolerance test<br>Decrease in Plasma TNFα ($p<0.05$)<br>No change in plasma IL-6<br>No change in muscle CD68 and MCP1 mRNAs |
| Rasouli<sup>53</sup><br>Yao-Borengasser<sup>28</sup> |  |  |  |  |  | Reduction in IMCL in type I fibres (Oil Red O staining, 34% reduction, $p=0.001$ vs. baseline)<br>No change in IMCL in type II fibres ($p=0.16$ vs. baseline)<br>No change in muscle SDH (a mitochondrial enzyme)<br>No change in mRNA levels of genes involved in muscle lipid oxidation (CPT1, PPAR-alpha, PPRAR-delta) |
| Bajaj<sup>37</sup> | Randomised<br>Controlled<br>Small sample | Type II Diabetes (n=15) | 45mg/day<br>Pioglitazone<br>(n=7) | Fenofibrate<br>200mg/day<br>(n=8) | 3 months | Increase in insulin-stimulated total-body glucose disposal ($p<0.005$), hyperinsulinemic-euglycemic clamp<br>Increase in AMPK activity following addition of pioglitazone to fenofibrate ($p<0.01$) |
| Glintborg<sup>32</sup> | Randomised<br>Placebo-controlled<br>Double-blind | Obese women of fertile age with polycystic ovary syndrome (n=24), age- and BMI-matched healthy women (n=14) | 30mg/day<br>Pioglitazone | Placebo | 16 weeks | Increase in insulin-stimulated Rd (36% increase, $p<0.001$), hyperinsulinemic-euglycemic clamp<br>Increase in insulin-stimulated GS (GSKB) activity (%I-form and % FV, pioglitazone group, $p<0.01$) |
| Hojlund<sup>54</sup> |  |  |  |  |  | Increase in Insulin-mediated AKT phosphorylation ($p<0.01$ vs pre-treatment insulin-mediated values)<br>No change in AMPK protein levels |
| Coletta<sup>33</sup> | Randomised<br>Double-blind<br>Parallel<br>study | Drug-naïve Type II<br>Diabetes (n =26) | 45mg/day<br>Pioglitazone<br>(n=14) | Aggressive<br>nutritional<br>therapy<br>(n=12) | 6 months | Increase in insulin-stimulated glucose disposal increased by 30% (p < 0.01),<br>hyperinsulinemic-euglycemic clamp<br>Increased AMPK phosphorylation (38%, p<0.05) |
| Ciaraldi<sup>39</sup> | Randomised | Polycystic Ovarian<br>Syndrome (n=20) and<br>healthy normal cycling<br>controls (n=12) | 45mg/day<br>Pioglitazone<br>(n= not<br>reported) | Placebo (n=<br>not<br>reported)) | 6 months | Increase in Rd (p<0.05), hyperinsulinemic-euglycemic clamp<br>Reduction in skeletal muscle CD15 (p<0.05)<br>Reduction in skeletal muscle CD68 (p = 0.052)<br>Reduction in skeletal muscle CXCR2 expression (receptor mediating neutrophil recruitment,<br>p=0.08)<br>Increase in expression of Iκβα (indicative of reduced inflammation, p< 0.05) |
| Bajpeyi<sup>34</sup> | Phase IV<br>trial<br>Randomised<br>Double-blind<br>Placebo-controlled | Obese, Type II Diabetes<br>(n=23) | 30mg/day<br>(first four<br>weeks), to<br>45mg/day if<br>HBA1c<br>>7.0%<br>(n=16, n=15<br>analysed) | Placebo<br>(n=8) | 12 weeks | Increase in insulin-mediated glucose disposal rate (p<0.001),hyperinsulinemic-euglycemic<br>clamp<br>Increase in change of respiratory quotient (p=0.001)<br>Decrease in IMCL content of gastrocnemius, tibialis anterior and soleus (p = 0.002, <0.001,<br>0.008 respectively)<br>No effect in resting ATP turnover (ATPase, p=0.56)<br>No effect on maximal ATP synthetic capacity (ATPmax, p=0.75) |
| Cusi<sup>35</sup><br>Bril | Randomised<br>Double-blind<br>Placebo-controlled | NASH plus pre-diabetes/Type II DM<br>(n=101) | 30mg/day,<br>up-titrated to<br>45mg/day<br>after 2<br>months | Placebo,<br>n=51 (n=42<br>completed<br>study) | 18 months | Increase in Rd (P<0.001), hyperinsulinemic-euglycemic clamp |
|  |  |  | N=50 (n=41 completed) |  |  |  |
| Tripathy<sup>38</sup> | Randomised Double-blind Placebo-controlled | Obese, Type II Diabetes (n=29) | 15mg/day (n=15, n=11 completed study) | Placebo (n=14, n=9 completed the study) | 6 months | Increase in whole body insulin sensitivity in pioglitazone group (p=0.04, p<0.01 between group change in M/I) (hyperinsulinemic-euglycemic clamp)<br>Reduction in Muscle TNF-α (30%, p=0.02)<br>Reduction in muscle TACE enzymatic activity (>80%, p=0.005, p=0.047 between-group change in TACE activity)<br>No change in plasma IL-6, FRK and MCP1 |
| Fiorentino<sup>36</sup> | Randomised Placebo-controlled | Obese, Type II diabetes (n=24) | 15mg/day (n=11) | Placebo (n=9) | 6 months | No change in Skeletal muscle protein levels of ODPX<br>Skeletal muscle protein levels of ATP5A, ETFA and CX6B1 increased [+33% (P ≤ 0.05), +60% (P ≤ 0.05) and + 33% (P = 0.01), respectively] |
| Thomas<sup>29</sup> | Randomised Double-blind Placebo-controlled | Male with familial combined hyperlipidemia (n=38, n=22 completed study) | 30mg/day Pioglitazone for four weeks, uptitrated to 45mg/day (n=10) | Placebo (n=12) | 16 weeks | Reduction in CRP (mg/dl (p=0.04 between-group change)<br>Increase in soleus IMCL (p=0.02 between-group difference)<br>No change in tibialis anterior IMCL (p=0.9) |
| Zanchi | Double-blind Randomised Cross-over | Non-diabetic, end-stage renal disease (haemodialysis (n=8), peritoneal dialysis (n=4)) | 45mg/day (n=10 analysed) | Placebo (n=9 analysed) | 16 weeks | No improvement in total glucose disposal, hyperinsulinemic-euglycemic clamp<br>Lower CRP (mg/L) at termination of intervention (p=0.07 between-group)<br>No effect on IL-6 |

Five studies examined muscle mass and the total number of patients studied was 174. Risk of bias assessment found four out of the five studies reporting changes in muscle mass to be at high risk of bias, in the main due to selective reporting of results (per-protocol only as opposed to intention-to-treat, figure 2). One study examined muscle compositional changes^23^. No studies examined physical function and symptoms. Only one study examined muscle function, randomising patients to a 16-week resistance training program with/without pioglitazone (15mg loading 30mg maintenance),where the combination led to greater Leg Press Power than either intervention alone.^24^

**Figure 2:**
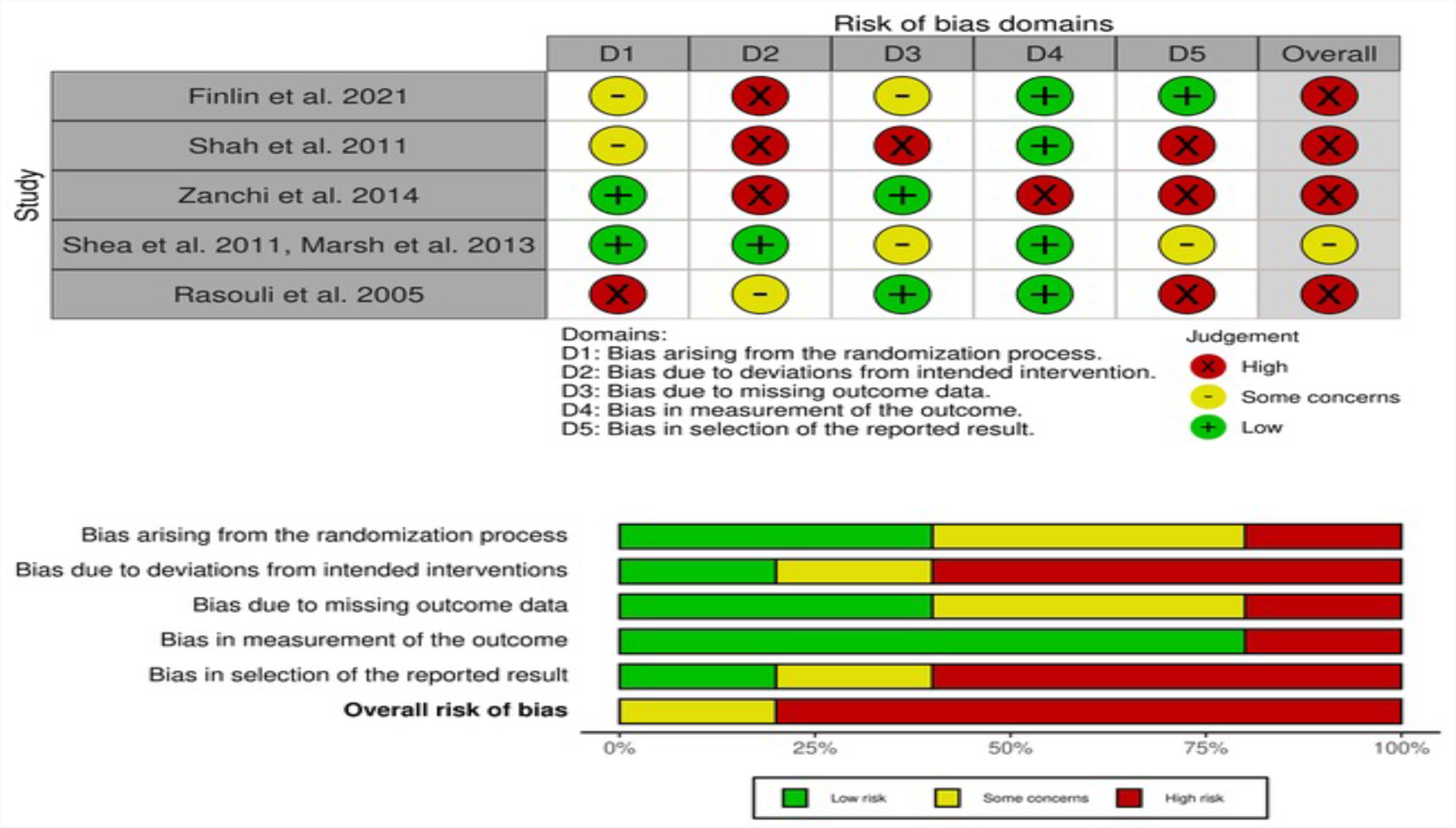
Risk of bias assessment for studies measures of muscle mass using Dual Energy X-ray Absorpiometry.

### Body composition

One study used Computer Tomography to examine muscle mass and body composition, and four used Dual Energy X-ray Absorptiometry (DEXA). In 44 patients, appendicular lean mass measured by CT was unaffected by low dose pioglitazone (15mg 3 weeks followed by 30mg for 3 weeks)^23^, and a 30mg/12 week regimen had no effect in 12 patients^25^. In 2 studies of 23 patients randomised to pioglitazone 45mg/10-16 weeks, an increase in lean body mass of 1.8-1.92kg was seen using Dual-energy X-ray absorbtiometry^26,27^. In a further small cross-over study of 12 patients with end-stage renal failure, an intervention of 45mg/16weeks did not alter lean body mass.^22^ Studies on body composition and muscle function are summarised in table 1.

### Intramyocellular lipid accumulation

From a total of four studies, three measured intramyocelluar lipid (IMCL) by Oil Red-O stain (n=23)^26,28^ and one used 1H-MRS (n=45)^29,30^. A single study of 19 patients where Hounsfield Units were calculated from Computer Tomography, did not show a difference in attenuation^23^. One study with maximum dose of 45mg/day pioglitazone showed reductions in IMCL in Gastrocnemius, Tibialis anterior and soleus^30^. A further study of 10 patients noted an increase in IMCL in the Soleus muscle of 35% but not in anterior tibialis^29^. One study using Oil Red O staining showed a 34% reduction in IMCL in type 1 fibres of Vastus Lateralis^26,28^.

### Peripheral insulin sensitivity

Seven studies examined peripheral insulin sensitivity, six using whole body glucose disappearance (R_d_) and one using Frequently sampled insulin-modified intravenous glucose tolerance test, totalling 273 patients. Four studies were judged to have a high risk of bias and no studies had low risk of bias (figure 3)^31-37^. R_d_ increased between 30-71% over treatment periods. Meta-analysis was conducted using mean change of R_d_ from baseline to endpoint and compared using a standardised mean difference (figure 5). The random effects model suggested an increase in mean change of muscle insulin sensitivity with pioglitazone (p<0.01), however there was high heterogeneity (I^2^ = 73%).

**Figure 3:**
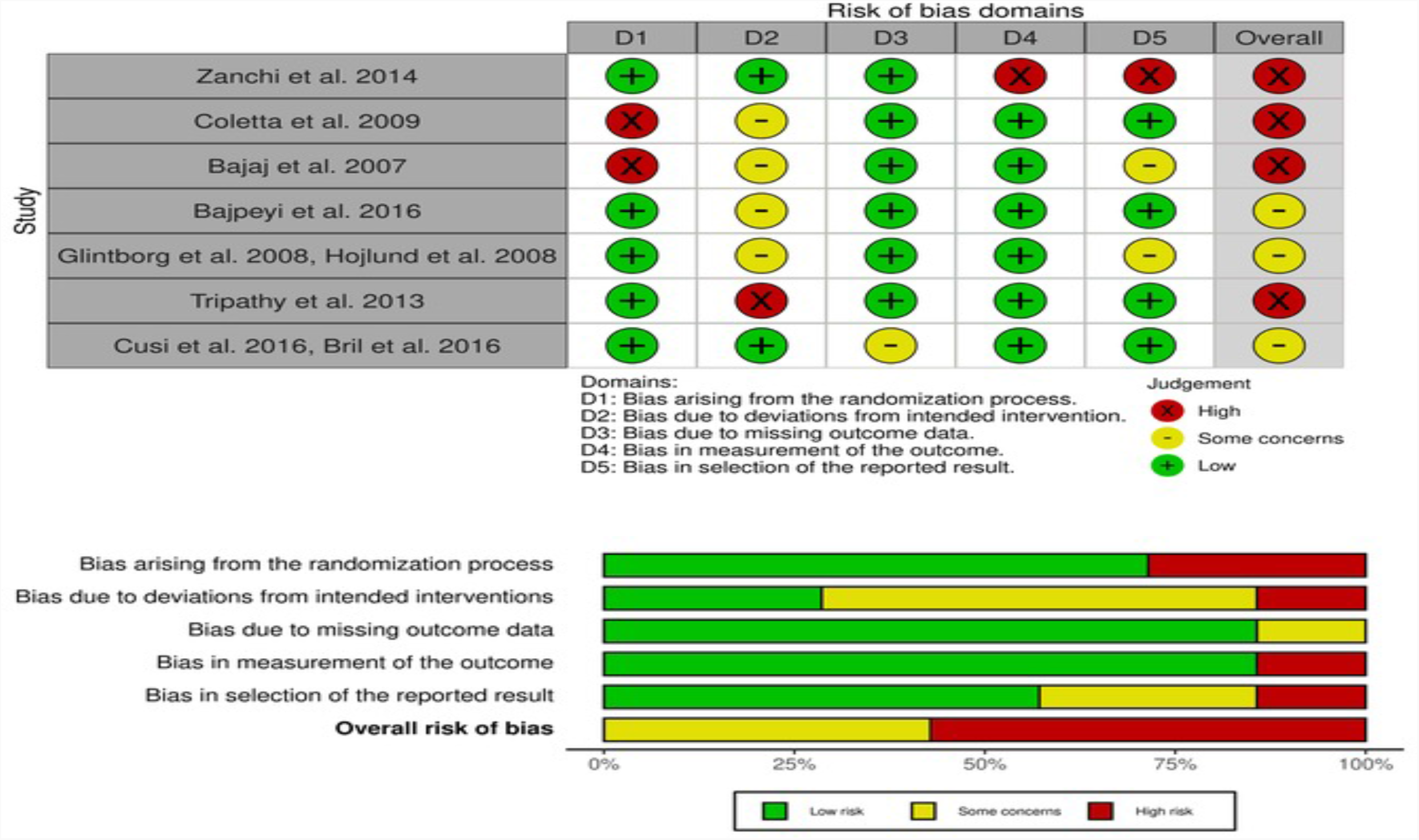
Risk of bias assessment for studies reporting changes in peripheral glucose disposal (R_d_)

**Figure 4:**
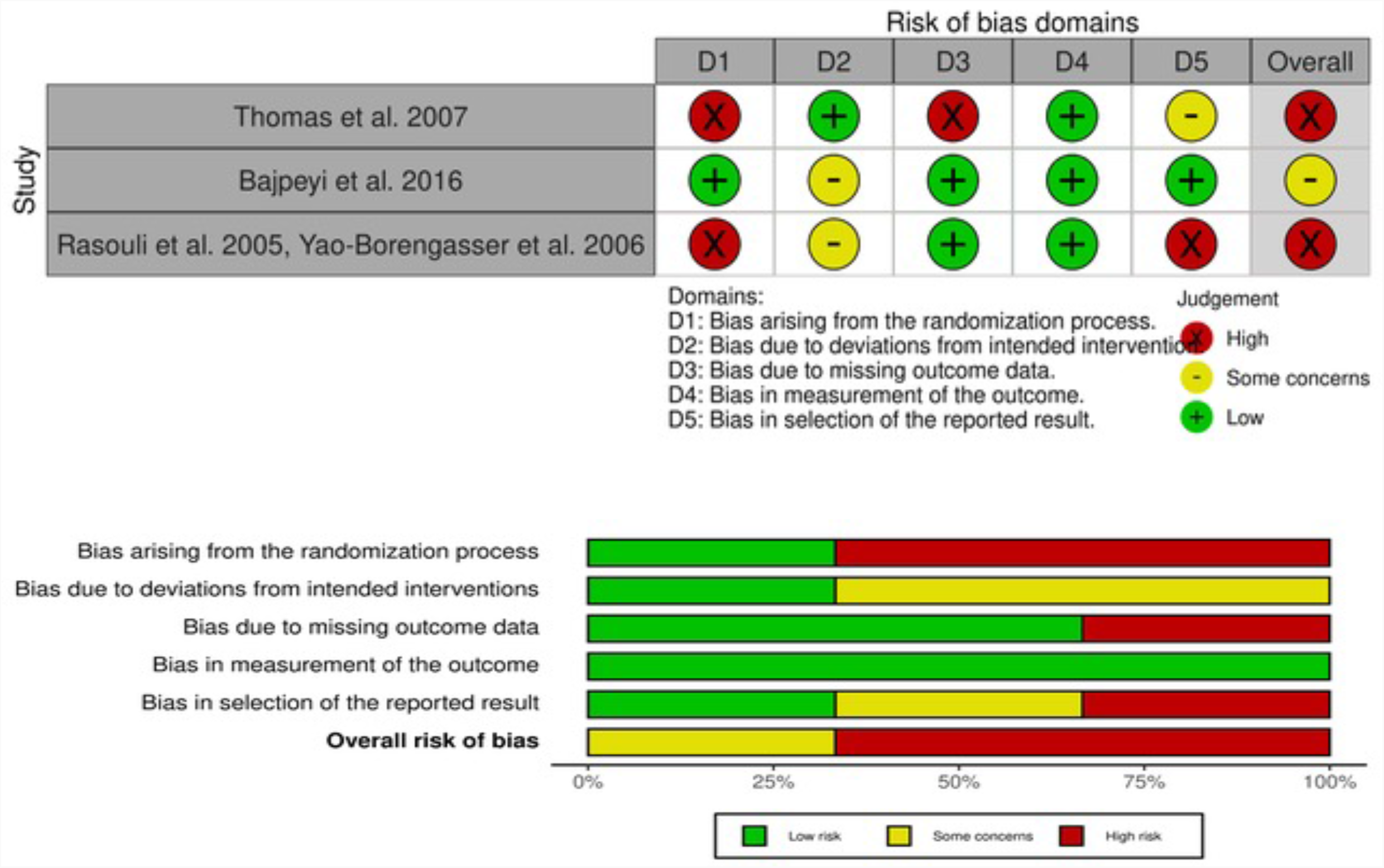
Risk of bias assessment for studies reporting changes in intramyocellular lipids.

**Figure 5:**
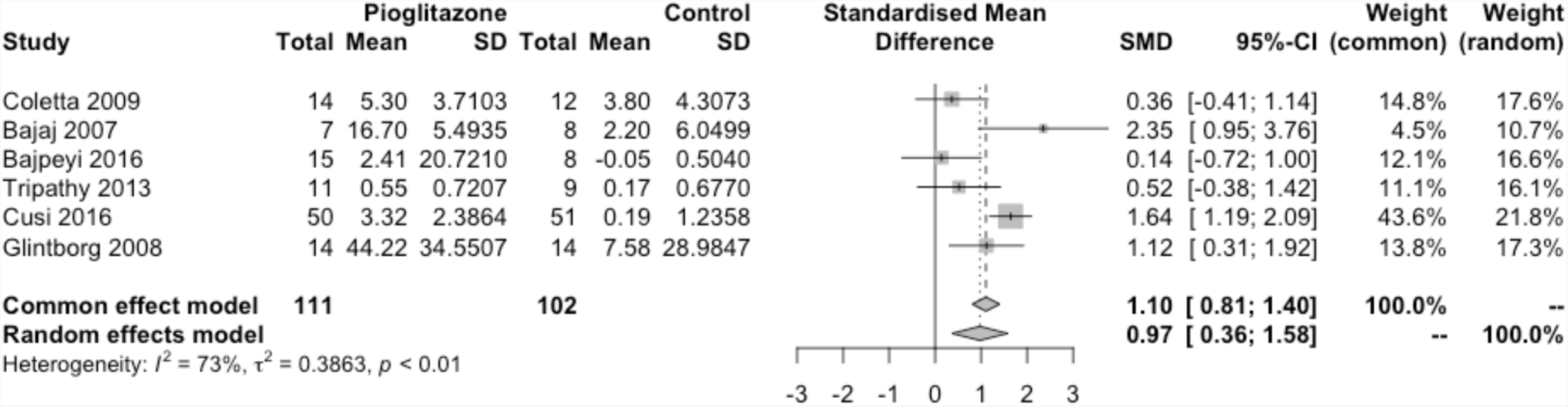
Forest plot of six studies reporting R_d_ demonstrated as a standardised mean difference. Mean change calculated as difference between endpoint mean and baseline mean presented. Standard deviation for mean change calculated using correlation coefficient (0.881).

**Figure 6:**
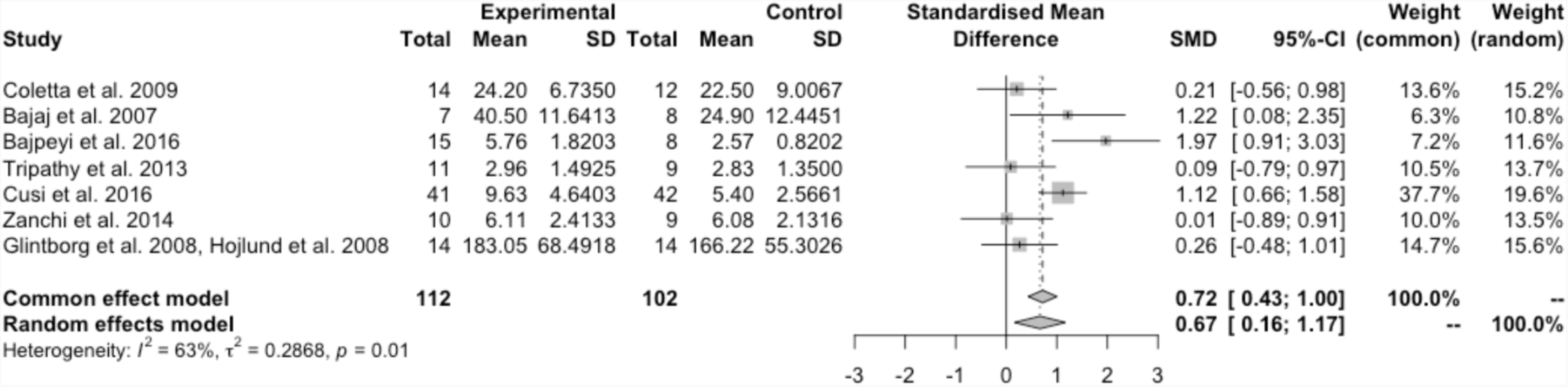
Forest plot of seven studies reporting R_d_. Analysis of end-point mean only using a standardised mean difference.

### Muscle gene expression, mitochondrial effects and markers of inflammation

One study noted no change in GLUT-4 concentration or phosphorylation of AS160, but did not examine GLUT-4 membrane translocation. One study of 24 patients demonstrated increased metabolic flexibility (ΔRQ between steady state and fasting RQ)^34^.

Tumour Necrosis Factor alpha (TNFα) concentrations decreased as a result of pioglitazone, both in plasma (19% n=53) and in muscle tissue (n=29), as did intramuscular TACE^31,38^. Further anti-inflammatory effects included decreases in circulating Interleukin 6, MCP-1 (n=29) and intramuscular Iκβα (n=32)^39^. One study examined the p44/p38JNk pathway and saw no discernable effect, and data on intramuscular CD68, CD15 and MCP-1 mRNA expression were inconclusive, potentially due to the low baseline intramuscular macrophage content in the subjects 2 studies (n=53)^31,39^.

Pioglitazone had conflicting effects on PAMPK/AMPK ratio^33,34,37^, but increased intramuscular markers of Oxidative Phosphorylation (n=55), mitochondrial biogenesis(PGC1α and PGC1β; n=26) and β-oxidation (n=29)^33,36^. One single study noted activation of the Insulin receptor pathway stimulating protein synthesis via Phosphoinositide 3-kinase (PI3K) and phosphorylation of Protein Kinase B (AKT) and Glycogen Storage Kinase-3-Beta (GSK3β)^32,40^. Studies examining secondary outcomes are summarised in table 2.

## Discussion

This systematic review and meta-analysis examined the effects of pioglitazone treatment on skeletal muscle metabolism and physiology. In 14 trials examined, only one study examined the effects of physical function measures demonstrating an improvement when combined with resistance exercise. The majority of studies measured muscle mass, which increased in studies that used higher doses of pioglitazone. Muscle insulin sensitivity increased in response to pioglitazone and intramyocellular lipid concentration and intramuscular inflammation decreased, with normalisation of metabolic function. In keeping with this, metabolic flexibility, oxidative phosphorylation and mitochondrial biogenesis markers normalised in response to pioglitazone.

In the main, muscle function is determined by muscle mass, which is maintained by a balance between muscle protein synthesis and muscle protein breakdown. Studies included in this systematic review rarely accounted for either resistance exercise or amino acid intake, with the exception of a single study. It would therefore not have been surprising to seen no change in muscle mass with pioglitazone therapy alone. The increase in muscle mass seen is likely to indicate increased anabolism following normalisation of metabolism and more efficient energy and protein utilisation. The Coefficient of Variation of DEXA is extremely low (0.3%) allowing some confidence in these measurements^41^. A key difference in acutely ill patients is that the clinical goal is not one of muscle gain, but one of maintenance, and these data suggest that normalisation of metabolism may be both possible and advantageous.

Insulin resistance is a hallmark of critical illness, and crucially, retains plasticity in response to interventions^42,43^. Glucose disposal (R_d_) measured during euglycaemic-hyperinsulinemic clamp with oral glucose tolerance testing reflects skeletal muscle insulin sensitivity, as endogenous hepatic glucose production is supressed^44,45^. Skeletal muscle insulin sensitivity increased in the majority of subjects given pioglitazone. While tight glucose control has not been shown to be feasible in a real-world setting, the fundamental importance of addressing insulin sensitivity remains important^46,47^. PPARγ agonism may be an appropriate alternative.

Critically ill patients have impaired beta-oxidation, leading to an increase in intramyocellular lipid concentrations^4,48^. A decrease in intramyocellular lipid content with pioglitazone therapy is likely to represent normalisation of beta-oxidation. The combination of returning substrate oxidative capacity in regards to carbohydrate and lipids to critically ill patients may occur with pioglitazone therapy, and address bioenergetic failure.

The pleiotrophic nature of PPARγ agonism precludes dissection if its anti-inflammatory effects from its effects on normalisation of substrate metabolism. Lipotoxicity from intramyocellular lipid accumulation and ceramide production may result in intramuscular inflammation^49^. Equally, systemic inflammation may result in decreased mitochondrial beta-oxidation, increasing lipid accumulation^4^. The causative pathway is likely to be bidirectional, and the decrease in both intramuscular and systemic inflammation (and specifically TNFα) lends further support for pioglitazone therapy.

Taking this data together, pioglitazone may counteract the acute metabolic effects of critical illness, allowing fat and carbohydrates delivered as part of clinical nutrition to be oxidised optimally, addressing bioenergetic failure. This in turn may allow muscle protein synthetic responses to normalise when amino acids or exercise are delivered, maintaining muscle mass and improving patient outcomes.

A limitation of this evidence synthesis is the heterogeneity of techniques used for assessment, and the lack of standardisation for concurrent exercise and nutrition. This results in conflicting data regarding bioenergetic sensing. The generalised concept of heterogeneous study design was partially addressed by the nature of the outcome measures chosen, allowing grouping of similar measures. A further limitation is the relatively small number of trials included. However pharmacological optimisation of clinical nutrition is still a relatively emerging discipline, and the depth of study data reflects this. The risk of bias of many of these studies was high, often due to per-protocol analysis of data as opposed to intention to treat. This is not uncommon in physiological studies, in attempting to maintain study integrity. However this violates the principle of randomisation, may inflate the effects of the intervention and limits external validity^50^. Data from this systematic review are hypothesis generating only, and this is an acceptable limitation to be addressed in future studies.

A major strength is the consistency in direction of data regarding normalisation of substrate utilisation, despite the small numbers. This provides weight to the need to test the effects of PPARγ agonism on optimising clinical nutrition utilisation to improve patient outcomes.

## Conclusions

Pioglitazone therapy increases skeletal muscle insulin sensitivity, decreases intramyocellular lipid accumulation and systemic and intramuscular inflammation. Where lean body mass was measured, this was seen to increase. Pioglitazone may be an adjunctive therapy to optimise clinical nutrition in acutely unwell patients.

## Data Availability

All data produced in the present study are available upon reasonable request to the authors

## Acknowledgements

The review authors are grateful to Dr D Glintborg for their co-operation in providing raw data for their trial included in this review.

## Notes

**Conflict of interest statement:** ZP has received honoraria for consultancy from Nestle, Nutriticia, Faraday Pharmaceuticals and Fresenius-Kabi, and speaker fees from Baxter, Fresenius-Kabi, Nutriticia and Nestle.

### Competing Interest Statement

ZP has received honoraria for consultancy from Nestle, Nutriticia, Faraday Pharmaceuticals and Fresenius-Kabi, and speaker fees from Baxter, Fresenius-Kabi, Nutriticia and Nestle.

### Funding Statement

This study did not receive any funding

