## Appendix 1 for "Pioglitazone for optimisation of clinical nutrition therapy in the critically ill patient: A Systematic Review"

**Database: Embase (Via Ovid), 1974 to January 23^rd^ 2022**

1. Pioglitazone/
2. (Pioglitazone OR Actos).ti,ab
3. OR/ 1-2
4. Physical Activity, Capacity and Performance/ OR Physical Performance/ OR Motor Performance/ OR (Physical Activity).ti,ab
5. Cardiorespiratory Fitness/ OR Fitness/ OR (Fitness).ti,ab
6. Functional Status/ OR Physical Mobility/ OR (Functional Performance).ti,ab
7. Exercise/ OR Exercise Test/ OR Walk Test/ OR Bicycle Exercise Test/ OR Cardiopulmonary Exercise Test/ OR Shuttle Walk Test/ OR Six Minute Walk Test/
8. (Exercise OR Exercise Test OR Walk Test OR Exertion OR Sit To Stand OR Timed Up and Go OR Functional Status).ti,ab
9. Physical Capacity/ OR Physical Tolerance/ OR Exercise Tolerance/ OR Physical Conditioning, Human/ OR Endurance/ OR Muscle Fatigue/ OR (Exercise Tolerance OR Endurance OR Fatigue OR Exercise Capacity).ti,ab
10. Muscle Strength / OR Hand Strength/ OR Grip Strength/ OR Pinch Strength/ OR (Musc* Strength OR Hand Strength OR Grip Strength OR Leg Press Power)ti,ab
11. Muscle Weakness/ OR (Musc* Weakness).ti,ab
12. Muscle/ OR Striated Muscle/ OR Muscle Activity/ OR Striated Muscle Cell/ OR Skeletal Muscle Cell/ OR Skeletal Muscle Contractility/ OR Skeletal Muscle Contraction/ OR Skeletal Muscle Enlargement/ OR Skeletal Muscle Fiber/ OR Striated Muscular Contractility/ OR Muscle Thickness/ OR (Musc* OR Muscle Cells OR Skeletal Muscle OR Striated Muscle OR Musc* Contraction).ti,ab
13. Frailty/ OR Muscle Atrophy/ OR Sarcopenia/ OR Paresis/ OR (Frailty OR Muscular Atrophy OR Sarcopenia OR Paresis) .ti,ab
14. Mitochondrion/ OR Muscle Mitochondrion/ OR Muscle Metabolism/ OR (Muscle Mitochondria OR Mitochondria).ti,ab
15. Muscle Protein/ OR (Muscle Proteins).ti,ab
16. Daily Life Activity/ OR Daily Life Activity Assessment/ OR Daily Living Activity/ OR (Activities Of Daily Living OR Performance Status OR Disability OR Mobility OR ADL).ti,ab
17. Nutritional Status/
18. Body Composition/ OR Body Distribution/ OR (Muscle Composition OR Musc* Thickness OR Muscle Gene Expression).ti,ab
19. Musculoskeletal System Inflammation/ OR Myositis/ OR (Musc* Inflammat* OR Musc* Adj5 Inflammat*).ti,ab
20. OR/4-19
21. Randomized Controlled Trial/
22. Controlled Clinical Trial/
23. Random$.ti,ab
24. Randomization/
25. Intermethod Comparison/
26. Placebo.ti,ab
27. (Compare OR Compared OR Comparison).ti
28. ((Evaluated OR Evaluate OR Evaluating OR Assessed OR Assess) AND (Compare OR Compared OR Comparing OR Comparison)).ab
29. (Open Adj Label).ti,ab
30. ((Double OR Single OR Doubly OR Singly) ADJ (Blind OR Blinded OR Blindly)).ti,ab
31. Double Blind Procedure/
32. Parallel Group$1 ti,ab
33. (Crossover OR Cross Over).ti,ab
34. ((Assign$ OR Match OR Matched OR Allocation) Adj5 (Alternate OR Group$1 OR Intervention$1 OR Patient$1 OR Subject$1 OR Participant$1)).ti,ab
35. (Assigned OR Allocated).ti,ab
36. (Controlled Adj7 (Study OR Design OR Trial)).ti,ab
37. (Volunteer OR Volunteers).ti,ab
38. Human Experiment/
39. Trial.ti
40. OR/21-39
41. (Random$ ADJ Sampl$ ADJ7 (Cross Section$ OR Questionnaire$1 OR Survey$ OR Database$1)).ti,ab NOT (Comparative Study/ OR Controlled Study/ OR Randomi?Ed Controlled.ti,ab OR Randomly Assigned.ti,ab)
42. Cross-Sectional Study/ NOT (Randomized Controlled Trial/ OR Controlled Clinical Study/ OR Controlled Study/ OR Randomi?Ed Controlled.ti,ab. OR Control Group$1.ti,ab)
43. (((Case ADJ Control$) AND Random$) Not Randomi?Ed Controlled).ti,ab
44. (Systematic Review Not (Trial OR Study)).ti
45. (Nonrandom$ NOT Random$).ti,ab
46. Random Field$.ti,ab
47. (Random Cluster ADJ3 Sampl$).ti,ab
48. (Review.ab AND Review.pt) Not Trial.ti
49. We Searched.ab AND (Review.ti OR Review.pt)
50. Update Review.ab
51. (Databases ADJ4 Searched).ab
52. (Rat OR Rats OR Mouse OR Mice OR Swine OR Porcine OR Murine OR Sheep OR Lambs OR Pigs OR Piglets OR Rabbit OR Rabbits OR Cat OR Cats OR Dog OR Dogs OR Cattle OR Bovine OR Monkey OR Monkeys OR Trout OR Marmoset$1).ti AND Animal Experiment/
53. Animal Experiment/ NOT (Human Experiment/ OR Human/)
54. OR/41-53
55. 40 NOT 54
56. AND/3, 20, 55

**Database: MEDLINE (via ovid), Ovid MEDLINE and Epub Ahead of Print, In-Process, In-Data-Review & other non-indexed citations from 1946 until January 23^rd^ 2022**

1. Pioglitazone/Exp
2. Pioglitazone.ti,ab
3. Actos.ti,ab
4. OR/1-3
5. Cardiorespiratory Fitness/Exp OR OR Physical Fitness/Exp OR OR Exercise/Exp OR OR Exercise Tolerance/Exp OR OR Physical Endurance/Exp OR OR Physical Exertion/Exp OR Fitness.ti,ab OR Exercise.ti,ab OR Exercise Tolerance.Ti,Ab OR Endurance.ti,ab. OR Exertion.ti,ab OR Exercise Capacity.ti,ab
6. Fatigue/Exp OR Muscle Fatigue/Exp OR Fatigue.ti,ab
7. Human Physical Conditioning/Exp
8. Exercise Test/Exp OR Walk Test/Exp OR Walk Test.ti,ab OR Exercise Test.Ti,Ab OR "Sit To Stand".ti,ab OR "Timed Up And Go".ti,ab OR Leg Press Power.ti,ab
9. Frailty/Exp OR Muscular Atrophy/Exp OR Sarcopenia/Exp OR Paresis/Exp OR Frailty.ti,ab OR Muscular Atrophy.ti,ab OR Sarcopenia.ti,ab OR Paresis.ti,ab
10. Muscle Strength/Exp OR Hand Strength/Exp OR Pinch Strength/Exp OR Musc* Strength.ti,ab OR Hand Strength.ti,ab OR Grip Strength.ti,ab
11. Muscle Weakness/Exp OR Musc* Weakness.ti,ab
12. Muscles/Exp OR Muscle Cells/Exp mOR Skeletal Muscle Fibers/Exp OR Skeletal Muscle/Exp OR Striated Muscle/Exp OR Musc$4.ti,ab OR Muscle Cells.ti,ab OR Skeletal Muscle.ti,ab OR Striated Muscle.ti,ab
13. Skeletal Muscle Enlargement/Exp OR Muscle Contraction/Exp OR Musc* Contraction.ti,ab OR Musc* Thickness.ti,ab
14. Muscle Mitochondria/Exp OR Muscle Proteins/Exp OR Mitochondria/Exp OR Muscle Mitochondria.ti,ab OR Mitochondria.ti,ab OR Muscle Proteins.ti,ab OR Muscle Gene Expression.ti,ab
15. Activities Of Daily Living/Exp OR Functional Status/Exp OR Karnofsky Performance Status/Exp OR Physical Functional Performance/Exp OR Functional Performance.ti,ab OR Activities Of Daily Living.ti,ab OR Functional Status.ti,ab OR Performance Status.ti,ab OR Disability.ti,ab OR Mobility.ti,ab OR ADL.ti,ab
16. Nutritional Status/Exp OR Body Constitution/Exp OR Muscle Composition.ti,ab
17. Muscular Diseases/Exp Or OR Musculoskeletal Abnormalities/Exp Or OR Myotoxicity/Exp Or OR Myositis/Exp OR Musc* Inflammat*.ti,ab OR (Musc* Adj5 Inflammat*).ti,ab
18. OR/5-17
19. Randomized Controlled Trial.pt
20. Controlled Clinical Trial.pt
21. Randomized.ab
22. Placebo.ab
23. Drug Therapy.fs
24. Randomly.ab
25. Trial.ab
26. Groups.ab
27. OR/19-26
28. Animals/Exp
29. Humans.SH
30. 28 NOT 29
31. 27 NOT 30
32. AND/4,18,31

**Database: CENTRAL, searched on January 23^rd^ 2022**

1. Pioglitazone/exp
2. pioglitazone ti,ab,kw
3. Actos
4. OR/1-3
5. Functional Performance ti,ab,kw OR Fitness ti,ab,kw OR Exercise ti,ab,kw OR Exercise Tolerance ti,ab,kw OR Endurance ti,ab,kw
6. Fatigue ti,ab,kw OR exercise test ti,ab,kw OR walk test ti,ab,kw OR frailty ti,ab,kw OR exertion ti,ab,kw
7. exercise capacity ti,ab,kw OR sit to stand ti,ab,kw OR timed up and go ti,ab,kw OR musc* strength ti,ab,kw OR hand strength ti,ab,kw
8. grip strength ti,ab,kw OR musc* weakness ti,ab,kw OR Musc* ti,ab,kw OR muscle cells ti,ab,kw OR skeletal muscle ti,ab,kw
9. muscle mitochondria ti,ab,kw OR mitochondria ti,ab,kw OR muscle proteins ti,ab,kw OR leg press power ti,ab,kw OR muscle composition ti,ab,kw
10. striated muscle ti,ab,kw OR musc* contraction ti,ab,kw OR muscular atrophy ti,ab,kw OR sarcopenia ti,ab,kw OR paresis ti,ab,kw
11. musc* thickness ti,ab,kw OR muscle gene expression ti,ab,kw OR activities of daily living ti,ab,kw OR functional status ti,ab,kw OR performance status ti,ab,kw
12. disability ti,ab,kw OR mobility ti,ab,kw OR ADL ti,ab,kw
13. musc* inflammat* ti,ab,kw OR (musc* ADJ5 inflammat*) ti,ab,kw
14. Exercise/exp OR Physical Functional Performance/exp OR Cardiorespiratory Fitness/exp OR Physical Fitness/exp OR Physical Endurance/exp
15. Exercise Test/exp OR Walk Test/exp
16. Exercise Tolerance/exp OR Physical Conditioning, Human/exp
17. Muscle Strength/exp OR Hand Strength/exp OR Pinch Strength/exp
18. Muscle Weakness/exp
19. Muscles/exp OR Muscle, Striated/exp OR Motor Activity/exp OR Cells/exp OR Skeletal Muscle Enlargement/exp OR Muscle Contraction/exp OR Muscle Fibers, Skeletal/exp
20. Muscle Proteins/exp
21. Muscle Fatigue/exp OR Frailty/exp OR Muscular Atrophy/exp OR Sarcopenia/exp OR Paresis/exp
22. Mitochondria/exp OR Mitochondria, Muscle/exp
23. Activities of Daily Living/exp OR Functional Status/exp
24. Nutritional Status/exp OR Body Composition/exp OR Constitution/exp
25. Myositis/exp
26. OR/5-25
27. AND/4, 26

**Database: CINAHL, searched on January 23^rd^ 2022**

1. Pioglitazone/exp
2. Pioglitazone ti,ab
3. Actos ti,ab
4. OR/exp1-3
5. Fitness ti,ab OR Exercise ti,ab OR Exercise/exp OR Exercise Tolerance ti,ab OR Endurance ti,ab OR Fatigue ti,ab OR Physical Fitness/exp OR Cardiorespiratory Fitness/exp OR Exertion ti,ab OR Exertion/exp OR Exercise Capacity ti,ab OR Muscle Fatigue/exp OR Physical Endurance/exp OR Exercise Tolerance/exp OR Aerobic Capacity/exp OR Fatigue/exp
6. Exercise Test ti,ab OR Walk Test ti,ab OR Sit To Stand ti,ab OR timed Up And Go ti,ab OR Leg Press Power ti,ab OR Exercise Test, Cardiopulmonary/exp OR Exercise Test/exp OR Exercise Test, Muscular/exp OR Grip Strength/exp
7. Muscular Atrophy ti,ab OR Sarcopenia ti,ab OR Paresis ti,ab OR Muscular Atrophy/exp OR Sarcopenia/exp
8. Physical Activity/exp OR Physical Performance/exp
9. Musc* Strength ti,ab OR Hand Strength ti,ab OR Grip Strength ti,ab OR Musc* Weakness ti,ab OR Musc* Contraction ti,ab OR Motor Activity/exp OR Muscle Strength/exp OR Muscle Weakness/exp OR Movement/exp
10. Musc* ti,ab OR Muscle Cells ti,ab OR Skeletal Muscle ti,ab OR Striated Muscle ti,ab OR Muscle Fibers/exp OR Muscle, Skeletal/exp OR Muscles/exp
11. Muscle Mitochondria ti,ab OR Mitochondria ti,ab OR Muscle Proteins ti,ab OR Muscle Gene expression ti,ab exp OR Muscle Proteins/exp OR Mitochondria/exp
12. Muscle Composition ti,ab OR Musc* Thickness ti,ab OR Nutritional Status/exp OR Body Composition/exp OR Adipose tissue Distribution/exp
13. Activities Of Daily Living ti,ab OR Functional Status ti,ab OR Functional Performance ti,ab OR Functional Status/exp OR Performance Status ti,ab OR Disability ti,ab OR Mobility ti,ab OR Physical Mobility/exp OR ADL ti,ab OR Karnofsky Performance Status/exp OR Barthel Index/exp OR Functional Assessment Inventory/exp OR Frailty ti,ab OR Frailty Syndrome/exp OR Activities Of Daily Living/exp
14. Myositis/exp OR Musculoskeletal abnormalities/exp OR Musc* Inflamm* ti,ab OR Musc* Inflammation ti,ab OR Musc* Adj5 Inflammat* ti,ab
15. OR/5-14
16. Randomized Controlled Trials/exp
17. Double-Blind Studies/exp
18. Single-Blind Studies/exp
19. Random Assignment/exp
20. Pretest-Posttest Design/exp
21. Cluster Sample/exp
22. (Randomised OR Randomized) .ti
23. Random*.ab
24. Trial .ti
25. (Sample Size/exp AND (Assigned OR Allocated OR Control).ab
26. Placebos/exp
27. Randomized Controlled Trial .pt
28. (Control W5 Group) .ab
29. (Crossover Design/exp OR Comparative Studies/exp)
30. (Cluster W3 RCT) .ab
31. OR/16-30
32. Animals/exp
33. Animal Studies/exp
34. Animal Model* .ti
35. OR/32-34
36. Human/exp
37. 35 NOT 36
38. 31 NOT 37
39. AND/4,15,38

**Database: ICTRP, searched on January 23^rd^ 2022**

1. Physical Activity OR Physical Performance OR Motor Performance OR Functional Performance
2. Cardiorespiratory Fitness OR Fitness OR Tolerance OR Functional Status
3. Physical Mobility OR Performance Status OR Disability OR Mobility
4. Exercise OR Exertion OR Exercise Test OR Walk Test OR Bicycle Exercise Test OR Cardiopulmonary Exercise Test OR Sit To Stand OR Timed Up And Go OR Leg Press Power
5. Physical Capacity OR Physical Tolerance OR Endurance
6. Muscle Strength OR Hand Strength OR Grip Strength OR Pinch Strength
7. Muscle Weakness OR Muscle Fatigue OR Muscular Fatigue OR Fatigue
8. Muscle OR Muscular OR Striated Muscle OR Skeletal Muscle OR Muscle Cells
9. Skeletal Muscle Contractility OR Skeletal Muscle Contraction OR Muscle Activity OR Muscular Activity OR Skeletal Muscle Enlargement OR Muscle Thickness
10. Frailty OR Sarcopenia OR Muscular Atrophy OR Paresis
11. Mitochondria OR Muscle Mitochondria OR Muscle Metabolism OR Muscle Proteins OR Muscle Gene Expression
12. Activities Of Daily Living OR ADL
13. Nutritional Status OR Body Composition OR Body Distribution OR Musc* Composition
14. Muscle Inflammation OR Muscular Inflammation OR Myositis
15. OR/1-14
16. Pioglitazone OR Actos
17. AND/15,16

**Database: Clinicaltrials.gov, searched on January 23^rd^ 2022**

1. Physical Activity OR Physical Performance OR Motor Performance OR Functional Performance
2. Cardiorespiratory Fitness OR Fitness OR Tolerance OR Functional Status
3. Physical Mobility OR Performance Status OR Disability OR Mobility
4. Exercise OR Exertion OR Exercise Test OR Walk Test OR Bicycle Exercise Test OR Cardiopulmonary Exercise Test OR Sit To Stand OR Timed Up And Go OR Leg Press Power
5. Physical Capacity OR Physical Tolerance OR Endurance
6. Muscle Strength OR Muscular Strength OR Hand Strength OR Grip Strength OR Pinch Strength
7. Muscular Weakness OR Muscle Weakness OR Muscle Fatigue OR Muscular Fatigue OR Fatigue
8. Muscle OR Muscular OR Striated Muscle OR Skeletal Muscle OR Muscle Cells
9. Skeletal Muscle Contractility OR Skeletal Muscle Contraction OR Muscle Activity OR Muscular Activity OR Skeletal Muscle Enlargement OR Muscle Thickness
10. Frailty OR Sarcopenia OR Muscle Atrophy OR Muscular Atrophy OR Paresis
11. Mitochondria OR Muscle Mitochondria OR Muscle Protein OR Muscle Metabolism OR Muscle Proteins OR Muscle Gene Expression
12. Activities Of Daily Living OR ADL
13. Nutritional Status OR Body Composition OR Body Distribution OR Muscle Composition
14. Muscle Inflammation OR Muscular Inflammation OR Myositis
15. OR/1-14
16. Pioglitazone OR Actos
17. AND,15,16
